# Remarkable variability in SARS-CoV-2 antibodies across Brazilian regions: nationwide serological household survey in 27 states

**DOI:** 10.1101/2020.05.30.20117531

**Authors:** Pedro C Hallal, Fernando P Hartwig, Bernardo L Horta, Gabriel D Victora, Mariângela F Silveira, Claudio J Struchiner, Luís Paulo Vidaleti, Nelson Arns Neumann, Lúcia C Pellanda, Odir A Dellagostin, Marcelo N Burattini, Ana M B Menezes, Fernando C Barros, Aluísio J D Barros, Cesar G Victora

## Abstract

Population based data on COVID-19 are essential for guiding public policies. We report on the first of a series of planned seroprevalence surveys relying upon on household probabilistic samples of 133 large sentinel cities in Brazil, including 25,025 participants from all 26 states and the Federal District. Seroprevalence of antibodies to SARS-CoV-2, assessed using a lateral flow rapid test, varied markedly across the country’s cities and regions, from below 1% in most cities in the South and Center-West regions to up to 25% in the city of Breves in the Amazon (North) region. Eleven of the 15 cities with the highest seroprevalence were located in the North, including the six cities with highest prevalence which were located along a 2,000 km stretch of the Amazon river. Overall seroprevalence for the 90 cities with sample size of 200 or greater was 1.4% (95% CI 1.3–1.6). Extrapolating this figure to the population of these cities, which represent 25% of the country’s population, led to an estimate of 760,000 cases, as compared to the 104,782 cases reported in official statistics. Seroprevalence did not vary significantly between infancy and age 79 years, but fell by approximately two-thirds after age 80 years. Prevalence was highest among indigenous people (3.7%) and lowest among whites (0.6%), a difference which was maintained when analyses were restricted to the North region, where most indigenous people live. Our results suggest that pandemic is highly heterogenous, with rapid escalation in Brazil’s North and Northeast, and slow progression in the South and Center-West regions.

---

Although the need for population-based data on COVID-19 is widely recognized,^1,2^ very few national surveys are available.^3^ Studies that have attempted to recruit population-based samples – as opposed to volunteers or convenience samples – have been recently carried out in a handful of countries. National studies using RT-PCR showed prevalence of 0.6% in Iceland^4^, 0.3% in Austria^5^, and 0.9% in Sweden.^6^ A national serological survey in Spain found a prevalence of 5.0%, ranging from less than 2% in some regions to 11% in Madrid.^7^ Studies among volunteers or convenience samples^4,8–10^ usually show higher prevalence than population-based studies.

The first case of COVID-19 in Brazil was reported on February 27 in the city of São Paulo, and by May 29, there are over 440,000 reported cases and 26,000 deaths. Based on reported events, eight out of the 27 Federation Units (26 states and the Federal District) present cumulative mortality rates above 10 per 100,000 inhabitants: four in the North (Amazonas, Pará, Amapá and Roraima), two in the Northeast (Ceará and Pernambuco) and two in the Southeast Region (Rio de Janeiro and São Paulo).

Three population-based studies are available from the Southern half of Brazil. Data were collected on April 11–13, April 25–27 and May 9–11 in nine large cities in Rio Grande do Sul state. Prevalence of antibodies using the Wondfo rapid test were 0.05%, 0.13% and 0.22% in the three waves, respectively.^11^ In the city of Ribeirão Preto (São Paulo state) prevalence was equal to 1.4% based on the same test^12^ and in six high-risk districts in São Paulo city prevalence was found to be 5.4% using a chemiluminescence immunoassay.^13^

Governmental response to the pandemic has been marked by controversy, with the country’s president opposing social distancing measures and minimizing the importance of COVID-19. Two consecutive Ministers of Health were either fired or resigned in less than one month due to opposition to the president’s stance. In contrast, most state governors and city mayors enforced closure of schools, shops and non-essential services, and more recently imposed lockdowns and compulsory use of masks in public spaces. Hospital services, particularly in the eight states mentioned above, are at the brink of collapse due to the high numbers of affected patients, particularly for intensive care.

From mid-April onwards, a number of mayors and governors have relaxed social distancing policies. While schools remain closed and public gatherings prohibited, industrial, commercial and services sectors are allowed to open daily for limited periods, while use of face masks continues to be enforced.

In spite of the staggering official statistics, a vast majority of cases are not being reported as testing is limited to severe illnesses, and there is also evidence that COVID-19 deaths are undercounted. In light of the present crisis, there is an urgent need for population-based data on the pandemic.

We conducted a nationwide seroprevalence survey in 133 sentinel cities in 26 Brazilian states and the Federal District (Figure 1). Cities were selected because they constitute the seats of the country’s intermediate regions and represent commercial and services hubs for surrounding urban and rural areas. Over an 8-day period (from to May 14 to 21), our field team visited a systematic sample of households in randomly selected census tracts. In each household, we administered a rapid SARS-CoV-2 antibody test to a randomly selected household member, followed by a brief questionnaire to collect sociodemographic data (see Online Methods for details).

**Figure 1.**
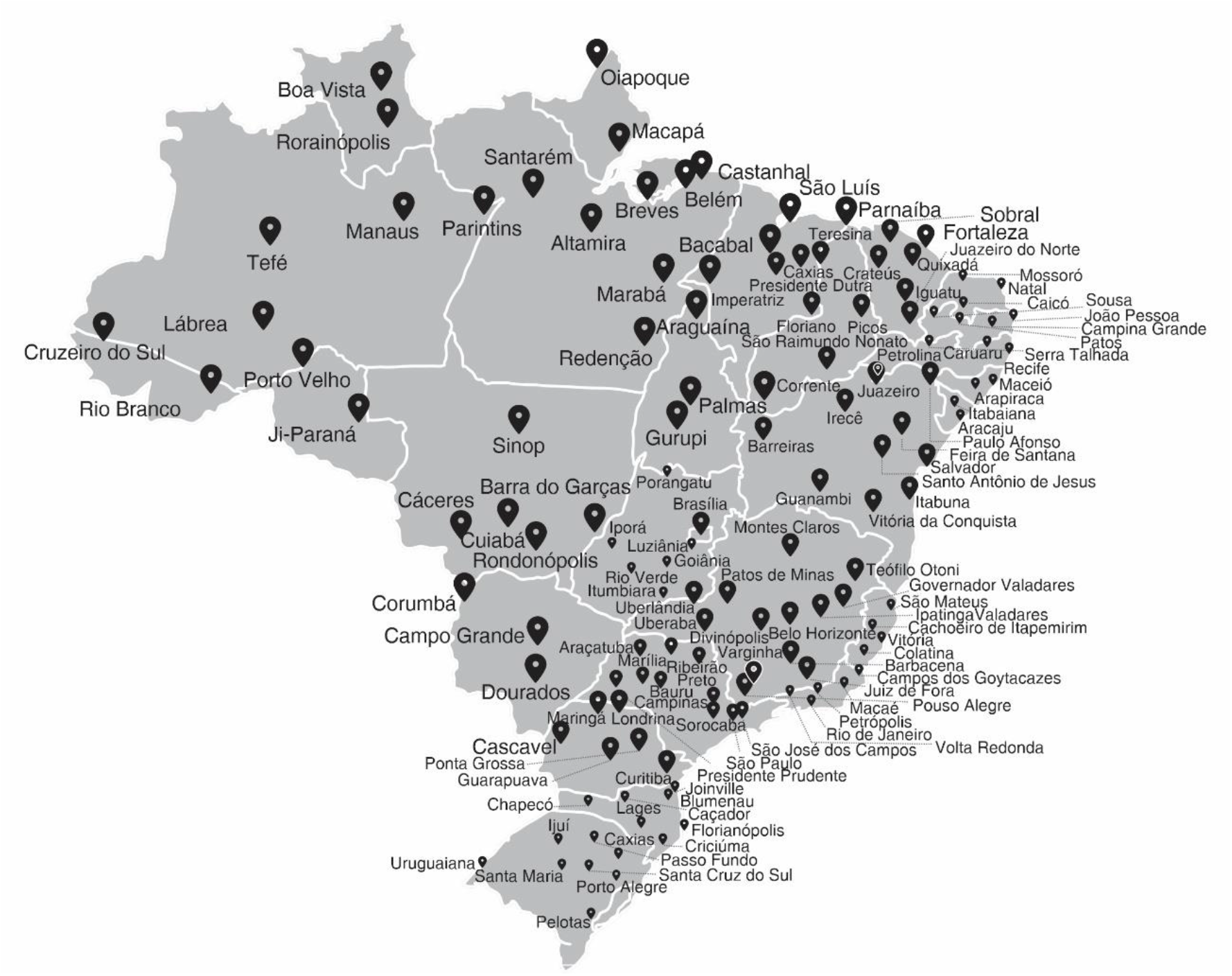
Location of the 133 sentinel cities.

The planned sample included 250 subjects in each of the 133 cities. It was not possible to conduct any interviews in one city; 250 individuals were tested in 46 cities, 200–249 in 44 cities, 100–199 in 14 cities, and 1–99 in 28. The total sample size was 25,025. The sample fell short of the planned number due to lockdown measures imposed in several cities with restrictions to mobility of the interviewers, and to lack of coordination between the Ministry of Health (which commissioned the study) and the city and state governments. These difficulties were compounded by the rapid spread of disinformation through social media characterizing the interviewers as swindlers, or of even being part of a plot to spread the virus. In 27 cities, interviewers were arrested and in eight cities the tests were destroyed by local police forces.

To reach a total of 25,025 interviews in the 133 selected cities, 46,011 attempts were made. Overall, 23% of the contacted households refused to participate and in another 23% residents were not available at the time of the visit, resulting in a response rate of 55%. Among the 90 cities where 200 or more interviews were completed, 18 had response rates of 80% or higher, 27 between 60% and 79%, and 45 below 60%. In terms of Brazil’s five regions, the average numbers of tests per city were 230 in the North, 156 in the Northeast, 177 in the Southeast, 242 in the South and 165 in the Center-West region. The national mean sample size by city was 188, or 75.2% of the target of 250 per city.

There are 5,565 cities in Brazil. We compared population sizes, reported COVID-19 cases and deaths by April 13, and the Human Development Index^14^ in three groups of cities: the 90 where it was possible to conduct 200 or more tests during the survey, the 43 cities included in the original sample where fewer than 200 subjects were tested, and the remaining 5,432 cities in the country (Supplementary Table 1). Cities with 200 or more tests tended to have larger populations and higher rates of reported cases and deaths than those with fewer than 200 tests, or the remaining cities in the country. The Human Development Index of the first two groups tended to be higher than in the third group of cities.

Table 1 shows the characteristics of individuals who provided blood samples. Compared to the whole Brazilian population, our sample of large sentinel cities included more individuals from the North and South regions, and fewer from the Southeast. Men and young people were underrepresented, as were those over the age of 50 years. The distribution in terms of skin color showed fewer individuals who reported being white, and larger percentages of black, Asians (yellow skin color) and indigenous people.

**Table 1.** Seroprevalence according to sociodemographic characteristics.

|  | Sample distribution |  | Brazil 2019 | Unadjusted |  | Adjusted for sample design and test validity |  |
| --- | --- | --- | --- | --- | --- | --- | --- |
|  | Number | % |  | Estimate | 95% CI | Estimate | 95% CI |
| **Region** |  |  |  | P<0.001 |  | P<0.001 |  |
| Northeast | 6552 | 26.2% | 27.2% | 0.7% | 0.5% - 0.9% | 0.8% | 0.5% - 1.1% |
| North | 5064 | 20.3% | 8.8% | 5.4% | 4.8% - 6.0% | 6.3% | 5.4% - 7.2% |
| Central-West | 2477 | 9.9% | 7.8% | 0.0% | 0.0% - 0.1% | 0.0% | NA% - NA% |
| Southeast | 5833 | 23.3% | 42.1% | 0.4% | 0.2% - 0.6% | 0.4% | 0.2% - 0.7% |
| South | 5069 | 20.3% | 14.3% | 0.1% | 0.1% - 0.3% | 0.1% | 0.0% - 0.6% |
| **Sex** |  |  |  | P=0.168 |  | P=0.225 |  |
| Female | 14452 | 57.8% | 51.7% | 1.3% | 1.1% - 1.5% | 1.5% | 1.2% - 1.8% |
| Male | 10543 | 42.2% | 48.3% | 1.5% | 1.3% - 1.8% | 1.7% | 1.4% - 2.0% |
| **Age (years)** |  |  |  | P=0.128 |  | P=0.323 |  |
| 0-4 | 430 | 1.7% | 7.2% | 1.4% | 0.5% - 3.0% | 1.6% | 0.6% - 3.5% |
| 5-9 | 682 | 2.7% | 7.0% | 1.2% | 0.5% - 2.3% | 1.3% | 0.5% - 2.9% |
| 10-19 | 2287 | 9.1% | 15.1% | 1.4% | 0.9% - 1.9% | 1.5% | 1.0% - 2.2% |
| 20-29 | 3866 | 15.5% | 16.5% | 1.4% | 1.1% - 1.8% | 1.6% | 1.2% - 2.1% |
| 30-39 | 3834 | 15.3% | 16.3% | 1.5% | 1.2% - 2.0% | 1.7% | 1.3% - 2.3% |
| 40-49 | 3975 | 15.9% | 13.5% | 1.6% | 1.2% - 2.0% | 1.8% | 1.4% - 2.3% |
| 50-59 | 4015 | 16.2% | 11.0% | 1.7% | 1.3% - 2.1% | 1.9% | 1.4% - 2.5% |
| 60-69 | 3381 | 13.5% | 7.5% | 1.0% | 0.7% - 1.4% | 1.1% | 0.7% - 1.6% |
| 70-79 | 1797 | 7.2% | 4.0% | 1.2% | 0.8% - 1.8% | 1.4% | 0.8% - 2.1% |
| 80+ | 728 | 2.9% | 2.0% | 0.5% | 0.1% - 1.4% | 0.6% | 0.1% - 1.6% |
| **Color/ethnicity** |  |  |  | P<0.001 |  | P<0.001 |  |
| White | 9493 | 38.7% | 45.2% | 0.6% | 0.5% - 0.8% | 0.7% | 0.5% - 0.9% |
| Brown | 11042 | 45.1% | 45.1% | 2.1% | 1.8% - 2.4% | 2.4% | 2.0% - 2.8% |
| Black | 2961 | 12.1% | 8.9% | 1.1% | 0.7% - 1.5% | 1.2% | 0.8% - 1.8% |
| Asian | 685 | 2.8% | 0.5% | 1.2% | 0.5% - 2.3% | 1.3% | 0.5% - 2.8% |
| Indigenous | 327 | 1.3% | 0.4% | 3.7% | 1.9% - 6.3% | 4.3% | 2.1% - 7.6% |

In total, there were 347 positive results in in 24,995 individuals with valid test results (1.39%). Prevalence results were corrected estimating a sensitivity of 84.8% based on four independent validation studies,^15^ and two different estimates of specificity, respectively 99.95% based upon our own early study in Southern Brazil,^11^ and 99.0% specificity as in validation studies using frozen sera.^15^ Prevalence results using the latter sets of estimates are presented in Supplementary Table 5; the largest difference in city prevalence levels using the two methods was 1.1 percent point.

Fifteen cities had prevalence above 2.0% (Supplementary Table 2): 11 in the North, two in the Northeast (Fortaleza with 8.9% and Recife with 3.4%) and two in the Southeast region (São Paulo with 3.3% and Rio de Janeiro with 2.4%). Four cities from the North showed prevalence above 10%: Breves (25.0%), Tefé (19.8%), Castanhal (15.5%), Belém (15.2%) and Manaus (12.7%). Except for São Paulo, where 212 individuals were tested, all other high-prevalence cities had samples of at least 240 subjects. The six cities with highest prevalence were all located along a 2,000 km stretch of the Amazon river, from Tefé in the central Amazon basin to Macapá, Belém, and Castanhal at the mouth of the river (Figure 2).

**Figure 2.**
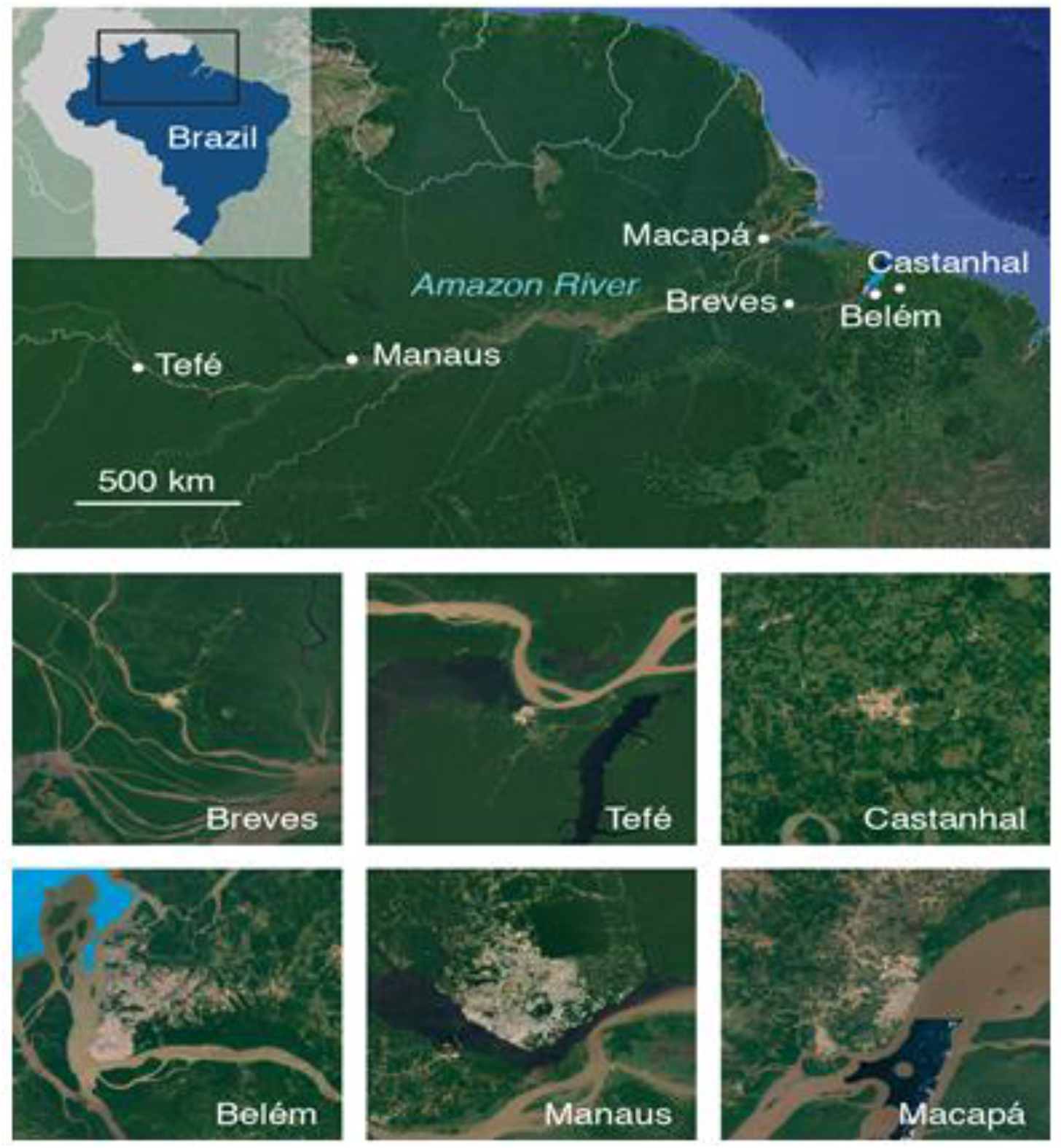
Six cities with the highest prevalence in the study. The small maps show areas of 50 × 50 km. line 518: Satellite images are from Google Earth.

Fifty-four (60%) of the 90 cities with at least 200 subjects had no positive tests, and 13 (14.4%) had only one positive test. Cities with zero cases represented 100% of those in the Center-West region (9 out of 9), 80% of those in the South (16 out of 20), 55% of those in the Southeast (11 out of 20), 55% of those in the Northeast (12 out of 22), and only 32% of those in the North (6 out of 19). When the index subject was positive, other household members were also tested; 21.6% of the families had at least another positive case. For families with at least two other members, this proportion was equal to 26.3%.

Figure 3 shows prevalence estimates as they relate to the officially reported COVID-19 cases and deaths in the country as of May 13, in the 90 cities with 200 or more tests. Due to the nature of antibody tests, very recent infections will not result in positive tests.^16^ This is partially offset by lags in reporting cases due to the time required for confirmation through RT-PCR testing, which according to local health authorities is of approximately two weeks. In addition, deaths are likely undercounted, and there may be a substantial time lag between infection and death, particularly in the case of prolonged hospitalization. In spite of these caveats, there were strong correlations between prevalence estimated by the survey and reported cases and deaths per population (Figure 3), with correlation coefficients of 0.66 and 0.85, respectively, both with P<0.0001. Our results regarding time trends were also consistent with officially reported cases and deaths (Figure 4).

**Figure 3.**
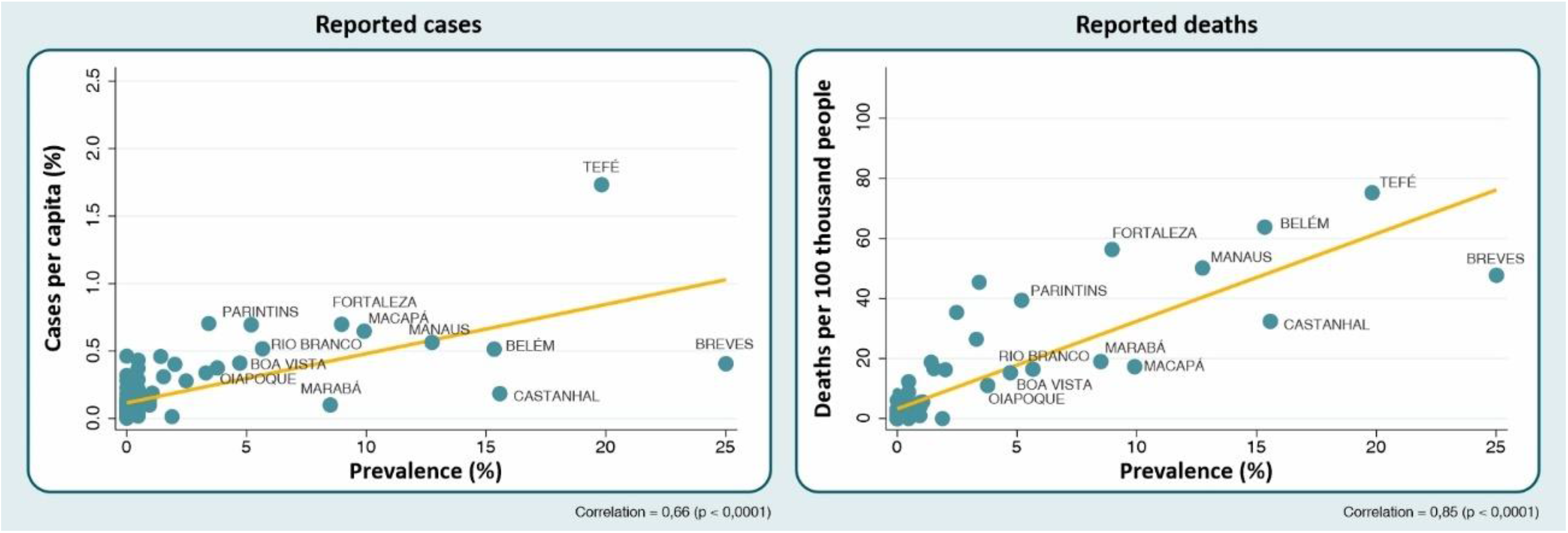
Scatter diagram for survey-based seroprevalence versus reported cases and deaths per population.

**Figure 4.**
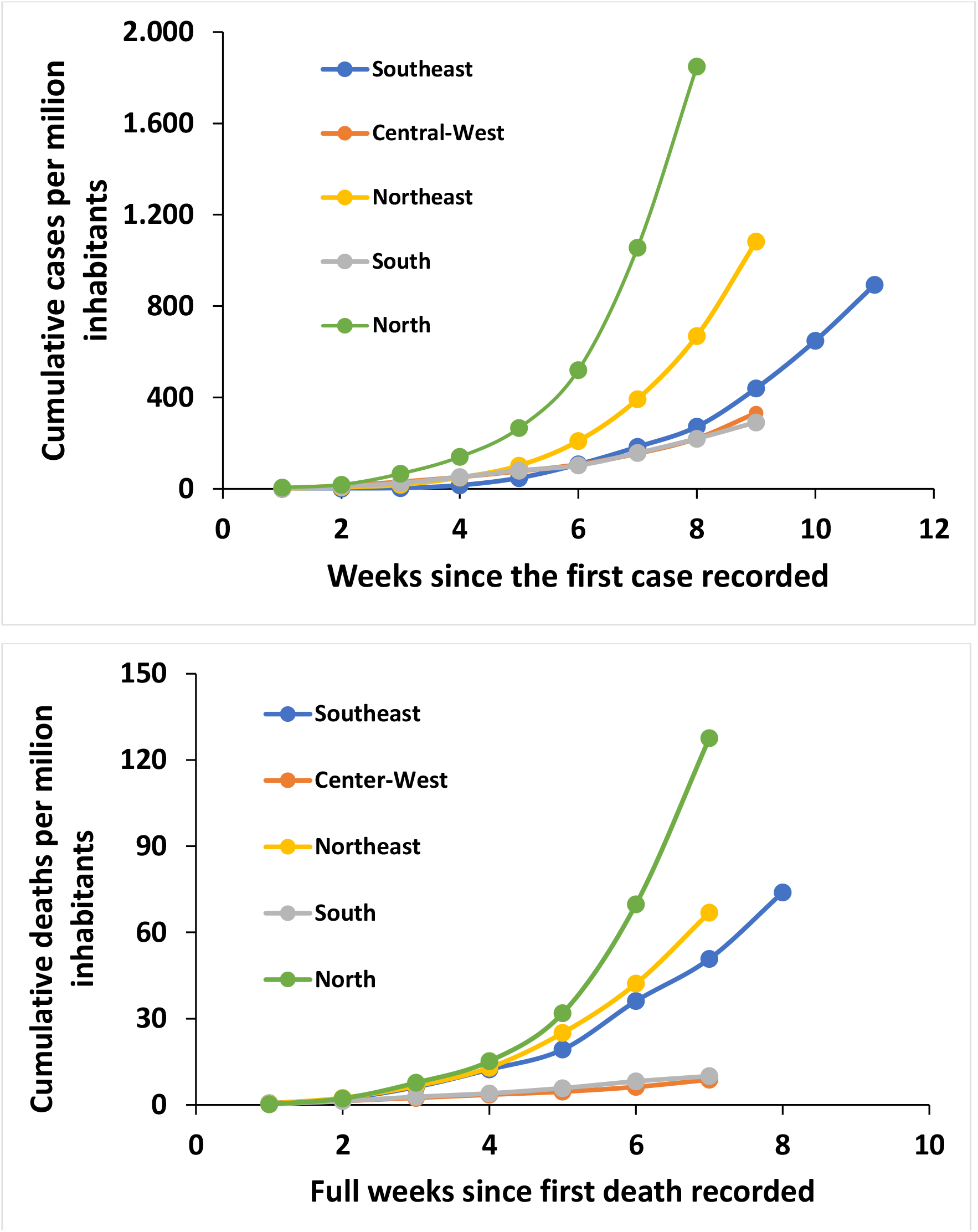
Time trends in reported cases and deaths between the start of the epidemic in each region and May 13, 2020 (source: https://covid.saude.gov.br)

We estimated the completeness of recording and infection-fatality rates for the 90 cities with samples of 200 or more tests. Taken together, these cities reported 104,782 cases by May 13, compared to our estimate of 760,000 based on the survey findings, a seven-fold difference – or, equivalently, 13.8% of underreporting. The ratio of deaths over estimated cases – the estimated infection-fatality rate – was 1.0% (7,640). Our estimates of underreporting at city level, for the 36 cities with non-zero prevalence, ranged from 0.4% to 57.3% of cases being officially reported, with estimated infection-fatality rates ranging from 0.0% to 2.4% (Supplementary Table 3).

Table 1 shows a breakdown of prevalence findings, resulting from two sets of individual-level analyses: unadjusted estimates, and estimates corrected for the test validity parameters and for the clustered nature of the sample. We focus the presentation of results on the second set. Individuals living in the North region had 6.3% prevalence, with the second highest prevalence observed in the Northeast (0.8%). No cases were observed in the Center-West region, and very few in the South.

Prevalence was similar among men and women. There was no significant difference according to age (P = 0.323) although individuals aged 80 or more years seem to be less frequently affected. Children aged 0–4 and 5–9 years were just as likely to display antibodies to SARS-CoV-2 as adults.

There were marked differences in prevalence according to ethnic group, ranging from 0.7% among whites to 4.3% in indigenous subjects. Because indigenous individuals in the sample were concentrated in the North region (35.5% of all indigenous in the national sample), and because most positive cases are also in the North, we carried out further analyses to verify whether the higher prevalence in indigenous individuals were due to confounding by region (Supplementary Table 4). In the national sample, the odds ratio for positive serology in indigenous individuals, compared to whites, was 5.89 (95%CI 2.99–10.66). Adjustment for region led to substantial reduction of the odds ratio to 2.26 (95%CI 1.13–4.17). Restriction of the analyses to the North region resulted in an odds ratio of 2.57 (95%CI 1.22–5.02), and further restriction to the cities in the North that are not state capitals produced an odds ratio of 2.45 (95%CI 1.00–5.40). P-values for the comparison of the five ethnic groups in the four sets of analyses described above were <0.001, 0.009, 0.003 and 0.030, respectively.

In terms of ethnicity, the second highest prevalence (2.4%) was found in the “brown” category. This is the most heterogeneous ethnic category in the country. A large nationwide genomic ancestry study^17^ showed that in the Northern city of Belém self-classified brown individuals had, on average, 69% European ancestry, followed by 21% Amerindian ancestry and 11% African ancestry, while in the South they had on average 44% European, 11% Amerindian and 45% African ancestries. Antibody prevalence in the brown group was 1.6-fold that of whites in the North, 1.3-fold in the Northeast and 0.9-fold in the Southeast.

To our knowledge this is the largest population-based study of prevalence of antibodies to SARS-CoV-2 in geographical scope, and the second largest – after the national survey in Spain^7^ – in terms of sample size. Possibly the most remarkable finding from our analyses was the cluster of high prevalence in six cities along the Amazon River. In the city of Breves, the prevalence of 25% appears to be the highest ever reported anywhere so far.^18^ This finding of high prevalence in a tropical region contradicts common wisdom that continents such as Africa may be protected against COVID-19 due to high ambient temperature.^19^

A possible explanation for this cluster is that long river trips (for example 8 hours from Belém to Breves, or 36 hours from Manaus to Tefé) offer the possibility of intense contagion in overcrowded boats where most passengers use hammocks for sleeping or resting in the decks (Figure 5). Except for road travel between Belém and Castanhal (67 km), all other transportation among the six cities is by river boat or – for a minority who can afford it – by plane. Unpublished analyses by Manaus city government showed an inverse association (Pearson’s r = 0.78; P<0.0001) between the daily number of boats leaving the capital to a given city and the number of days elapsed between the first reported case in Manaus and in the city of interest.^20^ The Ministry of Health started The reporting report daily numbers of cases on March 28, by when there were 105 cases in Manaus, 9 in Belém and 4 in Macapa – all state capitals – and one case in Castanhal which is close to Belém. The first case in Tefé was reported on April 10, and the first in Breves on April 20, or only three weeks before the reference date for the survey. high prevalence in the last two cities suggests that the epidemic was already well underway when the first case was reported.

**Figure 5.**
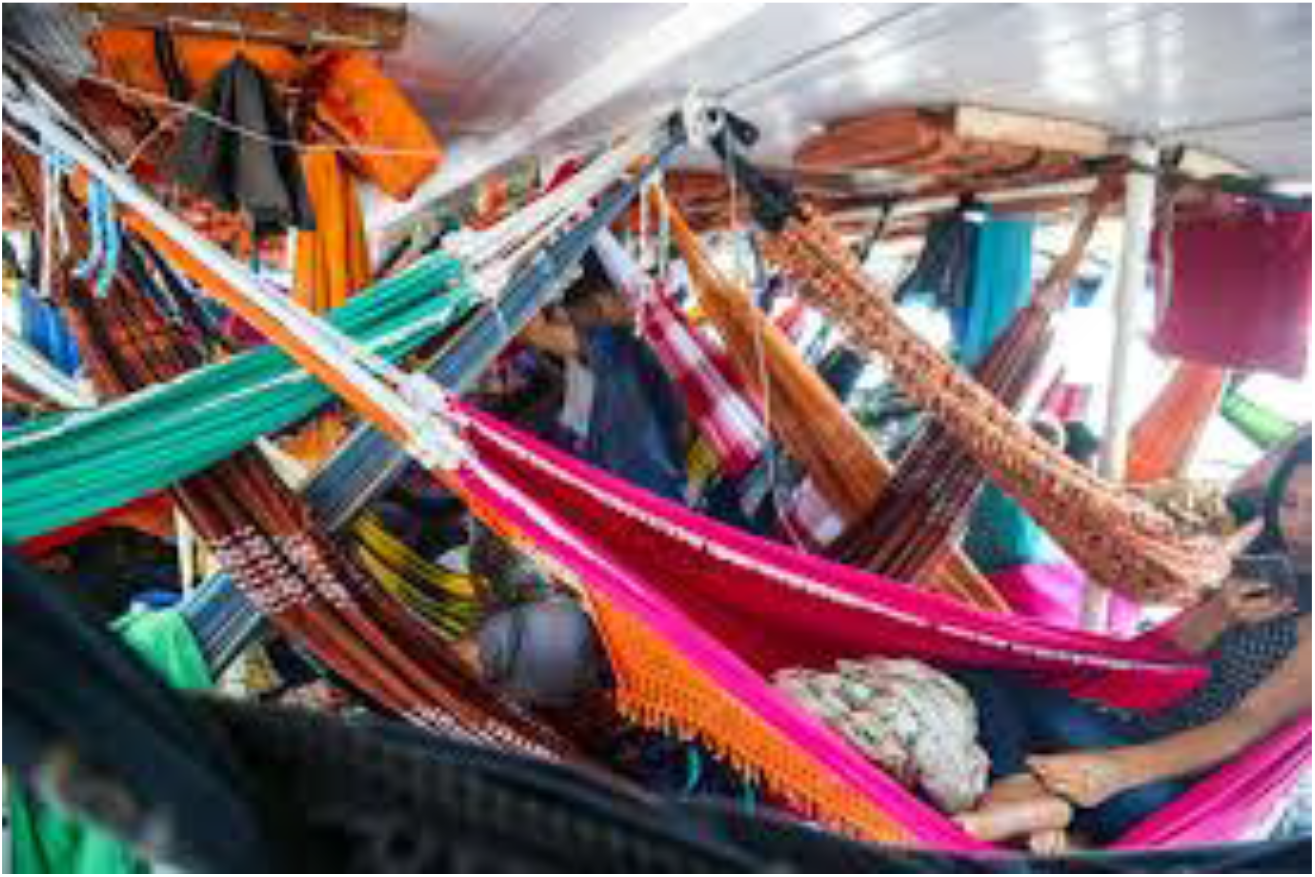
(illustrative). A crowded boat on the Amazon river.

Another possibility for high prevalence in the Amazon is that indigenous people, including those who self-classify as having “brown” skin color in the North region, may have higher susceptibility to SARS-CoV-2 infection due to genetic or sociocultural factors. According to the Articulation of Indigenous Peoples in Brazil,^21^ COVID-19 cases have already been detected in 71 different indigenous peoples around the country. Comorbidity with metabolic and cardiovascular diseases, which is increasing rapidly among native Brazilians^22^ would place them at increased risk of death due to COVID-19.Collection of biological samples in future studies may throw light on possible mechanisms.

We were only able to identify one population-based study of antibodies to SARS-CoV-2 in children. In Spain, prevalence was 1.1% among infants, 2.2% among children aged 1–4 years, and 3.0% at ages 5–9 years.^7^ All of these figures were lower than the overall study prevalence of 5%. In our study, young children displayed similar prevalence (1.3%) to that observed at older ages – for example, 1.4% both in 10–19 and 20–29-year-olds.

The limitations of our analyses include the restriction of the sample to sentinel cities that constitute regional hubs, which are larger, more developed and better equipped with health services than the country as a whole. Our survey response rate of 54.4% is similar to that in the Spanish survey (59.5%) and higher than achieved in national surveys in Iceland and Austria, both of which had response rates of about one third of the intended sample.^4,5^ In addition to the above-mentioned difficulties with local authorities and disinformation, it is known that many families have been moving away from large cities during the social isolation period, to stay with friends or relatives in small cities or rural areas. Our sample had fewer children than expected, which was probably due to their reluctance to undergo a finger prick when randomly selected within the household; in these cases, a second person was randomly selected and if that person also refused the household was replaced.

While concerns have been raised regarding the use of less than perfect serological tests for clinical decision-making and for defining individuals as immune to reinfection, use of such tests for population-based seroprevalence estimates is much less controversial, provided that sensitivity and specificity are sufficiently high and appropriately corrected for.^23,24^ The rapid lateral flow test used in our analysis (Wondfo SARS-CoV-2 antibody test) underwent four different validation studies, including one carried out by our own team. These studies placed the test’s sensitivity and specificity at 84.8% and 99.0%, respectively. This was the second best-performing lateral flow test out of 10 assessed by Whitman and colleagues.^25^ Given our concern that the test’s specificity in these four validation studies that relied on frozen sera is underestimated, we used our own specificity estimate from a statewide survey carried out in early April in nine cities with very few reported cases for COVID-19^15^ in the main analyses, while in the Supplementary Materials we also report results corrected for a specificity of 99.0% as calculated from the pooled validation studies (see Online Methods for more detail). The largest difference between the two sets of corrected estimates was 1.1 percent point.

Our serological results reflect the epidemic curve at a point in time that precedes sample collection by several days^16^. Reported cases and deaths are also affected by a variable time lag. Yet, our serological findings are consistent with reported deaths and cases (Figures 3 and 4). Our findings show on average one in seven cases were reported in these large cities, with above-average wealth and health services. However, this value was subject to large variation, with the highest-prevalence city of Breves reporting only 1.1% of infection. Our infection-fatality estimate based on reported deaths over survey-estimated cases was only 1.0%, but this statistic may be affected by underreporting of COVID-19 deaths.

We documented that the COVID-19 pandemic has affected the five regions of Brazil with widely different intensities. The highest prevalence was observed in a tropical area along the Amazon river. Individuals with indigenous ancestry were at particularly high risk. Young children were as likely to present antibodies as adults. This is the first wave of a national survey to be repeated every three weeks in order to monitor progress of the pandemic.

## Data Availability

Data will become publicly available upon request from the corresponding author 30 days after publication.

## Acknowledgments

We acknowledge the support from Instituto Serrapilheira, Pastoral da Criança, the Brazilian Collective Health Association (ABRASCO) and JBS’s initiative ‘Fazer o Bem Faz Bem’.

Pedro C Hallal, Fernando P Hartwig, Bernardo L Horta, Gabriel D Victora, Mariângela F Silveira, Claudio J Struchiner, Luis Paulo V Ruas, Lúcia C Pellanda, Odir A Dellagostin, Marcelo Burattini, Ana M B Menezes, Fernando C Barros, Aluísio J D Barros, and Cesar G Victora contributed to the conception and design of the work, to the acquisition, analysis, and interpretation of data and the draft of the manuscript. Nelson A Neumann contributed to the acquisition of data. All authors have approved the submitted version and have agreed to be personally accountable for the author’s own contributions and to ensure that questions related to the accuracy or integrity of any part of the work, even ones in which the author was not personally involved, are appropriately investigated, resolved, and the resolution documented in the literature.

## ONLINE METHODS

Brazil’s 27 federative units (26 states and the Brasilia Federal District) are divided by the National Institute of Geography and Statistics in 133 intermediary regions. The main city in each region was selected for the study (Figure 1). Further information on these sentinel cities is provided in the Supplementary Materials.

### Sampling

Using multistage sampling, we selected 25 census tracts with probability proportionate to size in each sentinel city, and 10 households at random in each tract. Using the data collection app, one individual was randomly selected from a listing of all household members completed at the beginning of the visit. Data collection took place from May 15–22, 2020. With 250 individuals per city, the margins of error for estimating prevalence figures of 2%, 5% and 10% are respectively 1.77, 2.70, and 3.79 percent points. At national level, the total desired sample of 33,250, the corresponding margins of error are 0.15, 0.24 and 0.33. In case the selected individual refused to provide a blood sample, a second household member was randomly selected. If this person also refused, the interviewers moved on to the next household to the right of the one that had been originally selected.

### Laboratory methods

Prevalence of antibodies was assessed with a rapid point-of-care test, the WONDFO SARS-CoV-2 Antibody Test (Wondfo Biotech Co., Guangzhou, China), using finger prick blood samples. This test detects immunoglobulins of both IgG and IgM isotypes specific to SARS-CoV-2 antigens in a lateral flow assay. Two drops of blood from a pinprick are sufficient to detect the presence of antibody. The assay reagent consists of colloidal gold particles coated with recombinant SARS-CoV-2 antigens. Following the introduction of the blood sample, reactive antibody:antigen:colloidal gold complexes, if present, are captured by antibodies against human IgM and IgG present on the on the “test” (T) line in the kit’s window, leading to the appearance of a dark-colored line. Samples without SARS-CoV-2-reactive antibodies will not lead to appearance of this line. Valid tests are identified by a positive control line (C) in the same window. If this control line is not visible, the test is deemed inconclusive, which is uncommon.

The rapid test underwent independent validation studies. According to the manufacturer, it has a sensitivity of 86.4% and specificity of 99.6% (https://en.wondfo.com.cn/product/wondfo-sars-cov-2-antibody-test-lateral-flow-method-2/). The tests were acquired by Brazilian Ministry of Health for population surveys and surveillance programs. A validation study carried out by the National Institute for Quality Control in Health (INCQS, Oswaldo Cruz Foundation, RJ, Brazil) showed a sensitivity of 100% and specificity of 98.7%. In an evaluation of 10 different lateral flow assays, Whitman and colleagues^25^ found that the Wondfo test was one of the two with the best performance, with sensitivity of 81.5% and specificity of 99.1%. Our own evaluation in Brazil found a sensitivity of 77.1% and specificity of 98.0%.^11^ By pooling the results from the four validation studies, weighted by sample sizes, sensitivity is estimated at 84.8% (95% CI 81.4%;87.8%) and specificity at 99.0% (95% CI 97.8%;99.7%).^11^

In early April 2020, our team conducted a household probability survey in nine cities in the state of Rio Grande do Sul (Nature Medicine, in press), when the pandemic was at a very early stage in the state. Of a total sample of 4,188 subjects there were only two positive results. We believe that this survey provides a better estimate of the test’s false-positive rate in the field, given that the other four studies relied on frozen samples for specificity estimation. Assuming that all cases in that survey were false-positives leads to a specificity rate of 99.95%. Whitman and colleagues, in their analyses of 10 lateral flow tests, observed “*moderate-to-strong positive bands in several pre-COVID-19 blood donor specimens, some of them positive by multiple assays, suggesting the possibility of non-specific binding of plasma proteins, non-specific antibodies, or cross-reactivity with other viruses*.”^25^ Our findings suggest the possibility that studies using frozen serum samples may have yielded higher false-positive rates than those associated with testing fingerprick blood. We therefore used as correction parameters in the main analyses a sensitivity of 84.8% and the 99.95% specificity derived from our previous population-based survey (Nature Medicine, in press). Analyses using the same sensitivity level and a specificity of 99.0% which is the weighted mean value of the first four validation studies are presented in the supplementary data.

### Data collection

Participants answered short questionnaires including sociodemographic information (sex, age, schooling, skin color and household assets), COVID-19-related symptoms, use of health services, compliance with social distancing measures and use of masks. Due to the presence of widespread miscegenation, the official Brazilian classification of ethnicity recognizes five groups, based on the question: “What is your race or color?” The five response options are “white”, “brown” (“pardo” in Portuguese), “black”, “yellow” and “indigenous”. Interviewers are instructed to check the “yellow” option when the respondent mentions being of Asian descent, and “indigenous” when any of the multiple first nations are mentioned. This system is endorsed by the Afro-descendants movement, which advocates for disaggregation of all national statistics to raise their visibility.^26^

Field workers used tablets or smartphones to record the full interviews, register all answers, and photograph the test results. All positive or inconclusive tests were read by a second observer, as well as 20% of the negative tests. If the index subject in a household had a positive result, all other family members were invited to be tested.

### Ethical approval and data availability

Interviewers were tested and found to be negative for the virus, and were provided with individual protection equipment that was discarded after visiting each home. Ethical approval was obtained from the Brazilian’s National Ethics Committee (process number CAAE 30721520.7.1001.5313), with written informed consent from all participants. Positive cases were reported to the municipal COVID-19 surveillance systems. Data will become publicly available upon request from the corresponding author 30 days after publication.

### Data analyses

The survey data was analyzed using two strategies. The first consists of treating the survey as if it were a simple random sample, using the exact binomial method to calculate confidence intervals and the likelihood ratio test (implemented as logistic regression) to compare the prevalence among sociodemographic groups. In the second strategy, we accounted for both the sampling design of the survey and corrected for the test validity, as described in detail in the Supplement. Hypothesis testing was performed using Cochran’s Q heterogeneity test implemented as fixed effects meta-regression. All analyses were performed using R version 3.6.1.^1^ The “survey” package^1,2^ was used to account for the sampling design. Meta-regression was implemented using the “metafor” package.^4^Further information on the analytical approach is available in the Supplementary Materials.

